# Designing the ASPIRE-SSI study: a multicenter, observational, prospective cohort study to assess the incidence and risk factors of surgical site and bloodstream infections caused by *Staphylococcus aureus* in Europe

**DOI:** 10.1101/2020.07.08.20148791

**Authors:** Darren P.R. Troeman, Susanne Weber, Derek Hazard, Martin Wolkewitz, Leen Timbermount, Tuba Vilken, Stephan Harbarth, Omar Ali, Frangiscos Sifakis, Jan A.J.W. Kluytmans

**Author notes:** **Corresponding author:** Darren P.R. Troeman, Julius Center for Health Sciences and Primary care, Utrecht Medical Center Utrecht, Postal address: Huispost nr. STR 6.131, P.O. Box 85500, 3508 GA Utrecht, The Netherlands.

## Abstract

**Background:** There is a continuing need for in-depth and updated knowledge about the epidemiology of surgical site infections (SSIs) caused by *Staphylococcus aureus* to support the development of effective preventive interventions. The ASPIRE-SSI study aims primarily to determine the incidence of *S. aureus* SSIs and postoperative bloodstream infections (BSIs) in Europe and to assess their association with patient-related, pathogen-related, and contextual risk factors.

**Methods:** ASPIRE-SSI is a prospective, multicenter, observational cohort study primarily assessing the incidence of and risk factors for *S. aureus* SSI and postoperative BSI in Europe. Five thousand adult surgical patients (of which two-thirds will be *S. aureus* carriers and one-third non-carriers) undergoing several types of surgical procedures in sites located across Europe were enrolled in the study. Data and specimens were collected from these subjects who were followed for up to 90 days following surgery to assess study outcomes.

Using advanced survival analyses and regression techniques (including competing risks models), we will determine event-specific and sub-distribution hazards to assess the independent associations of these study outcomes with risk factors. Additionally, a risk prediction model will be derived to quantify the risk of developing SSI or BSI due to *S. aureus*.

**Discussion:** Despite the challenges, this study will provide important and contemporary information about the epidemiology of SSI and BSI (and other infections) caused by *S. aureus* in the current surgical population in Europe, thereby supporting the development of effective preventive interventions.

**Trial registration:** clinicaltrial.gov number **NCT02935244**

## INTRODUCTION

Surgical patients are at risk of acquiring healthcare-associated infections (HAIs) (1). Studies show that up to 10% of these patients acquire at least one type of HAI during their hospital stay (2, 3). A substantial proportion of HAIs developed among surgical patients are surgical site infections (SSIs) (2, 4), which are associated with considerable morbidity and mortality, and increased healthcare costs (5, 6). *Staphylococcus aureus*, which is a commensal in approximately 20-30% of the healthy human population (7, 8), is one of the most common and important pathogens that cause SSIs (4, 9). Nasal carriage of *S. aureus* has been identified as an important risk factor for *S. aureus* infection (7, 8).

Despite the current standard of care, availability of antimicrobial therapies, and use of infection control measures, infections caused by *S. aureus* remain a serious health condition in high-risk patient populations, including surgical patients (10). In addition, the emergence of antibiotic-resistant *S. aureus* strains has increased the burden of staphylococcal disease (11). Being able to accurately identify patients in an early stage who bear a disproportionate risk of *S. aureus* infection may be an important approach in improving the prevention and management of these infections (12, 13), as well as allowing novel interventions to target such high-risk patients (14). To effectively study and implement such therapies, a current, in-depth understanding of the epidemiology and natural history of *S. aureus* infections in this population is critical. Prospective studies, such as this one, will enable the assessment of risk factors by which the subsets of patients that may benefit most from these interventions can be identified.

As such, the aim of this study is to systematically assess the patient-related, pathogen-related, and contextual factors on the incidence of *S. aureus* infections, particularly *S. aureus* SSI and postoperative bloodstream infection (BSI), in the current population of surgical patients in Europe. In addition, we aim to identify the patient subgroups that bear a disproportionate high risk of developing an *S. aureus* infection after surgery.

## METHODS

### Study design

The ASPIRE-SSI study, which is an acronym for *<u>A</u>dvanced understanding of <u>S</u>ta<u>p</u>hylococcus aureus infections in Europ<u>e</u> – <u>S</u>urgical <u>S</u>ite Infections*, is a prospective, observational, multicenter cohort study. It is part of the COMBACTE consortium (COMBatting AntibiotiC resisTance in Europe) (15), which aims to expedite the development of novel antimicrobial treatments and diagnostic tests in the fight against antibiotic resistance.

Two study populations can be distinguished within the ASPIRE-SSI study; a source population and a study cohort population. The study cohort population is nested within the source population, meaning that all data and study specimens that are collected for the study cohort population is in addition to the data that is collected for the source population. Sites in countries located across the four European sub-regions, as described by the United Nations (16), enrolled patients for this study. Patient enrollment started in December 2016 and ended in January 2020. Approvals from the Research Ethics Committees/Institutional Review Boards in the different countries/sites were obtained prior to patient enrollment at any site. A scheme of the study procedures can be found in the online supplemental material of this article (Supplemental Material: Tables 1 and 2).

### Study objectives

The main aims of this study are:

- To systematically assess the incidence of *S. aureus* SSI (composite of superficial, deep, and organ/space *S. aureus* SSI) and BSI up to 90 day following surgery, and determine its independent association with patient-related, pathogen-related and contextual factors in the current population of surgical patients in Europe.
- To develop risk prediction models to quantify the risk of developing a *S. aureus* SSI (composite of superficial, deep, and organ/space *S. aureus* SSI) and BSI following surgery in the current population of surgical patients in Europe.

A complete list of study objectives can be found on Clinicaltrials.gov, under study identifier **NCT02935244**, or in the online supplemental material of this article (Supplemental Material: Study Objectives).

### Eligibility criteria

#### Inclusion criteria

The following **inclusion criteria** were met by every subject that participated in the ASPIRE-SSI study:

1. The subject was 18 years of age or older.
2. The subject underwent one of the surgical procedures^*^ included in the study. The surgical procedure was planned or unplanned.
3. The subject underwent *S. aureus* screening of the nose, throat and perineum within 30 days prior to surgery to assess *S. aureus* colonization, and based on the preoperative *S. aureus* colonization status of the subject, the subject qualified for enrollment in the study cohort population. Only in the case of emergency surgery when it was not possible to screen the three body regions, could subjects qualified for enrollment in the study based on *S. aureus* screening of at least two body regions, including the nose.
4. Written informed consent was obtained prior to enrollment in the study cohort population.

*The surgical procedures included in this study were: craniotomy, laminectomy, spinal fusion, central artery reconstructive surgery, peripheral artery bypass surgery, mastectomy, open cardiac surgery, implantable cardioverter defibrillator (ICD) implantation, emergency surgery, hip prosthesis surgery and knee prosthesis surgery. The rationale for including these procedures in this study can be found in the Discussion’s section of this research paper.

#### Exclusion criteria

Potential subjects who met any of the following **exclusion criteria** were excluded from participation in the study:

1. Parallel participation in any preventative anti-Staphylococcal intervention study.
2. An active SSI diagnosis as the reason for undergoing surgery.
3. Not able to comply with study procedures and follow-up based on investigator judgment.

### Patient recruitment and study populations

Adult patients (i.e. 18 of age or older) undergoing one of the surgical procedures included in the study were potentially eligible for inclusion in the study and comprised the **target population** of the study. The local study teams or treating specialist (or delegate), as per local regulations, approached these patients to obtain informed consent. If a patient was unable to give informed consent, an authorized surrogate of the patient was approached for consent on the patient’s behalf, as per local regulations. The patients who consented were screened for *S. aureus* colonization in the nose, throat and perineum within 30 days prior to surgery. These subjects comprised the **source population** of the study. These subjects could subsequently be included in the **study cohort population** using an enrichment approach that was based on the preoperative *S. aureus* colonization status. The preoperative *S. aureus* screening procedure was standardized across participating sites and laboratories for consistency of screening results. We provided each participating site with screening swabs and swabbing instructions, including instructions on how to store the swabs and transport the swabs to the local laboratories. Also, each participating laboratory received the same plating materials and processing instructions, including instructions on how to culture the samples and report the screening results. The collected screening swabs were analyzed immediately by semi-quantitative culture on chromogenic agar plates (Colorex agar, bioTRADING Benelux) at the local laboratories to determine the preoperative *S. aureus* colonization status of the subjects.

We used the following enrichment approach to enroll study subjects from the source population in the study cohort: for every two *S. aureus* colonized subjects we included one non-colonized subject, provided that they underwent surgery. Only subjects with evaluable results for all three preoperative screening samples could be classified as either *S. aureus* colonized (at least one result positive for *S. aureus*) or non-colonized (three negative results for *S. aureus*) and could therefore be enrolled in the study cohort population. An exception to this rule was made for subjects undergoing emergency surgery, as the collection of all three screening samples could be challenging in the emergency setting. For these subjects, evaluable results of at least two swabs, including the nasal swab, were needed for the determination of the preoperative *S. aureus* colonization status of the subject. For this study, we defined emergency surgery as an unplanned surgical procedure that was performed on a subject whose clinical acuity was assessed as requiring surgery within 72 hours after hospital admission.

To ensure that subjects were included in the study cohort population before they developed the first signs of postoperative infection, we instructed the local study teams to enroll subjects in the study cohort within 72 hours after the index surgery. To ensure that an evaluable number of subjects was enrolled in the study cohort for each of the protocol-defined surgical procedures, we also stratified enrolment according to surgical procedure. Once the subjects were enrolled in the study cohort population, they were followed for 90 days following their surgery to assess study outcomes. An overview of the study procedures per study population can be found in the online supplemental material of this article (Supplemental Material: Tables 1 and 2).

### Study outcomes

The primary outcome of this study evaluates the incidence of *S. aureus* SSI (composite of superficial, deep, and organ/space SSI) up to 90 days following surgery. The main secondary outcomes include the incidence of serious *S. aureus* SSI (i.e. deep and organ space SSI); *S. aureus* bloodstream infections (BSIs) after surgery; as well as all postsurgical *S. aureus* infections. *S. aureus* strains involved in colonization and infection will also be characterized and the role of anti-*S. aureus* antibodies as potential biomarkers for *S. aureus* infection will be explored. In addition, we will develop a prediction model to quantify the risk of acquiring a *S. aureus* SSI or BSI using a composite score of independent risk factors. The study definitions for the various types of infections, including SSIs, are based on the widely-used CDC definitions (17, 18). A complete list of study outcomes can be found on Clinicaltrials.gov, under study identifier **NCT02935244**, or in the online supplemental material of this article (Supplemental Material: Study outcomes).

### Site selection

Sites could only be selected for this study if they had at least one corresponding microbiology laboratory that fulfilled the study requirements with respect to e.g. sample processing and storage, and that could be selected for this study. The sites were selected from lists of hospitals and corresponding microbiology laboratories that are part of the COMBACTE consortium (CLIN-Net and LAB-Net networks respectively) (15) and that meet certain quality and capability requirements. A total of 34 sites with corresponding microbiology laboratories in 10 different European countries were selected to take part in this study.

The sites and laboratories were selected by a Site Selection Committee (SSC) comprised of representatives from the University Medical Center Utrecht, the University of Antwerp, and MedImmune/AstraZeneca. The selection process was based on pre-defined criteria described in a Site Selection Plan. These criteria were based on the capability of the hospital staff to access and recruit the target population for the study and to collect follow-up data and specimens from study participants; the experience of the hospital staff with conducting clinical studies; the capability of the local microbiology laboratory; and the anticipated timelines for regulatory approvals and contracting negotiations. The assessment of these criteria was carried out by feasibility questionnaires completed by the sites and laboratories. Only sites that fulfilled all study requirements and that were considered fully capable of taking part in the study by all voting SSC members based on the feasibility data, were selected for the study.

To ensure that the study cohort population would be representative of the European continent (even outside the European Union), a pan-European approach was undertaken when assessing and selecting participating sites for this study. Sites were selected such that factors like geography, background antibiotic resistance prevalence, and number and type of surgical procedures were adequately balanced between the four European sub-regions (16). At least two countries from the Northern, Southern, Eastern and Western European region were included. To prevent delays in enrolment and ensure study completion within expected timelines, back-up sites were selected to replace or supplement primary selected sites that could withdraw from participation or that would have lower than expected recruitment rates.

### Statistical methods

#### Sample size calculation

The sample size for the study cohort population was based on the expected incidence precision of *S. aureus* SSI. Assuming an incidence of *S. aureus* SSI of approximately 1.5% in the target population (19), a prevalence of *S. aureus* colonization in the target population of 20% (7) and an eightfold (20) increased risk of developing an *S. aureus* SSI for subjects colonized with *S. aureus* compared with non-colonized subjects, we expected an *S. aureus* SSI incidence of 5% for *S. aureus* colonized subjects and 0.6% for non-colonized subjects. Furthermore, in the calculation of the sample size we also took into account that the study population should reflect the pan-European surgical population and that all eligible types of surgical procedures for inclusion in the study would be represented in sufficient numbers (see section “eligibility criteria” for an overview of the eligible surgical procedures). Based on this, we calculated that approximately 500 subjects needed to be enrolled per surgical procedure (except for spinal fusion and laminectomy, as these two surgical procedures will together include 500 patients), of which two-thirds would be *S. aureus* colonized. In total, we planned to enroll approximately 5000 patients in the study cohort. With this sample size, we expected an incidence estimate of *S. aureus* SSI of approximately 3% within each surgical procedure, with an incidence precision of 1.5% (95% confidence interval 1.5-4.5, using normal approximation). To achieve this sample size, we assumed that approximately 15.000 to 20.000 patients would need to be screened for assessment of preoperative *S. aureus* colonization status.

#### Planned analysis

The primary analysis will evaluate the cumulative incidence of *S. aureus* SSI up to 90 days following surgery. Estimates of cumulative incidence of serious *S. aureus* SSI, *S. aureus* BSI, other postoperative *S. aureus* infections, and all-cause SSI will also be calculated. In a secondary analysis, the incidence density of *S. aureus* SSI and postoperative mortality up to 90 days following surgery will be estimated.

Competing risk methodology will be used to estimate the incidence functions (21). Death without *S. aureus* SSI will be considered a competing event for *S. aureus* SSI. We will apply adapted Cox regression models for event-specific and sub-distribution hazards of SSIs to determine independent associations between outcomes and risk factors. To account for clustering of data (e.g. patients undergoing the same surgical procedure, patients in the same country) and random effects by sites, shared frailty methodology, stratification or robust variance will be applied. Hazard ratios with 95% confidence intervals will be calculated univariately and selected for multivariate model using established Akaike’s information criterion for model selection (22, 23). Missing values will be accounted for using multiple imputation (24).

The risk prediction model will be derived using a logistic regression model and candidate predictors identified through the risk factor analysis. For variable selection, a backward selection methodology will be applied. The overall model performance will be assessed by measuring the explained variation and by determining the calibration and discrimination of the model. The final model will be internally validated by bootstrapping (12). In a secondary analysis we will take the *time-to-event* of interest (e.g. *S. aureus* SSI) into account, and consider death without the event of interest as a possible competing event. In order to account for the matching procedure in the risk prediction model, we will incorporate weights into the model. Weights will be calculated using data collected from the source population. This will ensure that the population used for the derivation of the risk prediction model is representative of the target population.

### Quality assurance

The participating sites collected data from the study subjects and entered this data into a web-based electronic data capture system specifically built for this study. All participating study subjects received a subject ID. The information linking this subject ID to the subject’s medical information at the site was kept at a secure location at the participating site.

This study was also monitored. The monitoring by external auditors included 100% source data verification (SDV) for 10% of the total number of subjects enrolled in the study cohort at each participating site, including the SDV of the first three enrolled study cohort subjects (with a maximum of 15 study cohort subjects per participating hospital). In addition, each study cohort subject with an infection diagnosis and/or positive culture results underwent monitoring by at least one medical monitor to assure that the reported data was accurate, complete and in agreement with the study definitions for infections.

## DISCUSSION

This manuscript describes the study design and objectives of the ASPIRE-SSI study, a prospective observational study that aims to evaluate the occurrence of SSI and other postoperative infections caused by *S. aureus* in surgical patients in Europe and to identify subgroups of patients at high risk of these infections. During the conception of this study we made certain choices that require further discussion.

### Surgical procedures

The surgical procedures for this study were selected based on our current knowledge of surgical procedures that are widely performed and that are associated with a substantial burden of postoperative *S. aureus* infections (18). This makes them potential targets for future anti-staphylococcal preventive interventions. We assumed the following when selecting the surgical procedures for this study: (1) the main risk factor for developing an SSI is the surgical procedure itself from the time of incision to wound closure, (2) there is no *S. aureus* strain specificity for certain surgical procedures, and (3) the pathophysiology of and patient- and procedure-related risk factors for *S. aureus* SSI infections following these surgical procedures would be similar across similar surgeries (i.e. the results can be extrapolated to similar surgical procedures).

### Enrichment strategy and representativeness of study cohort population

Nasal *S. aureus* colonization has already been identified as an important risk factor for developing infections with *S. aureus*, particularly in the surgical setting. However, studies have shown that the prevalence of *S. aureus* colonization in the community is only about 20 to 30% (7, 8). For this reason, we enriched the study cohort population with *S. aureus* colonized subjects. We did this by enrolling *S. aureus* colonized and non-colonized subjects in a 1:2 ratio in the study cohort, thereby increasing the prevalence of *S. aureus* colonization in the study cohort population to 67%. By doing this, we expected to increase the number of *S. aureus* SSIs events, which in turn would allow for better incidence determination, risk prediction and biomarker estimations. Additionally, by including non-colonized subjects sequentially after at least two colonized subjects, we expected to be able to assess differences between *S. aureus* colonized and non-colonized subjects and ensure a similar temporal distribution of enrolled *S. aureus* colonized and non-colonized subjects. We are convinced that this will increase the robustness of the study results.

Because only a selection of screened subjects was enrolled in the study cohort, one could argue whether the study cohort population was representative of the larger surgical population. This is especially important when the data collected is used for risk prediction. We tried our best to ensure that the study cohort population would be a good representation of the target population through our study design. However, to be able to explore this further, we collected completely anonymized baseline data on certain factors (e.g. age and sex) from the entire target population. This data will be used to compare the distribution of these factors between the study cohort and the broader target population. By doing this, we will be able to assess the external validity of the study cohort population and ascertain potential selection bias between study participants and non-participants. In addition, we will use the target population data to derive weights that will be incorporated in the risk prediction models as a way to account for the matching procedure.

### Preoperative *S. aureus* colonization

Subjects from the source population were enrolled in the study cohort based on their preoperative colonization status. However, the determination of preoperative colonization status was based only on a one-time measurement of swabs taken from the nose, throat and perineum. Studies have shown that approximately 20% of the healthy human population is persistent *S. aureus* carrier, 30% is intermittent *S. aureus* carrier, and 50% is persistent non-carrier of *S. aureus* in the nose (7). Thus, it is highly probable that the *S. aureus* colonized patients identified in this study were a mixture of persistent and intermittent carriers, whereas the non-carriers of *S. aureus* were a mixture of persistent non-carriers and intermittent carriers.

Persistent carriers are known to carry higher loads of *S. aureus* and to shed bacteria more heavily than intermittent carriers (25, 26). On the contrary, carriers with lower loads of *S. aureus* may not be easily identified as carriers of *S. aureus* when screening swabs are directly plated onto culture media without prior cultivation in growth media (as is done in this study). Because of this, patients with lower loads of *S. aureus* may wrongly be classified as non-carriers, which could lead to misclassification. While this could be the case, the method used in this study to screen patients matched normal clinical practice, which makes the results more generalizable. Also, patients with very low loads of *S. aureus* colonization (i.e. colonization is only evident after enriching the culture sample prior to plating) are probably not at a higher risk of infection with *S. aureus* (26). Consequently, we did not anticipate that this would lead to a noteworthy bias. However, by doing semi-quantitative culture of the screening swabs (as is done in this study), we will get an estimation of the bacterial load of *S. aureus* colonization of each enrolled *S. aureus* carrier. This will hopefully allow us to assess whether our assumptions are corroborated by the study data.

Additionally, patients could be colonized with *S. aureus* at several body regions or extra-nasally only, and screening patients by swabbing only the nose could wrongly identify a patient as being non-carrier. By swabbing three body regions for *S. aureus* colonization, we have increased our chances of identifying all *S. aureus* carriers.

Another potential source of misclassification could be that in some countries and sites it was mandatory to decolonize patients for *S. aureus* colonization (targeted or universal) prior to certain surgical procedures, in particular in the presence of previously known MRSA colonization (12). If decolonization was performed after a patient was found to be colonized with *S. aureus* following preoperative *S. aureus* screening, this patient would have been included in the study cohort as *S. aureus* colonized, whereas the patient would have been decolonized before undergoing the surgical procedure. In such cases, the risk of patients developing a postoperative infection with *S. aureus* would be lower than if they would have not received any decolonization treatment (27, 28). If we would not correct for this, it could lead to erroneous conclusions. Therefore, we collect data on antibiotic usage, including data on preoperative decolonization treatment, and will be able to account for this in the analysis.

### Complexity of study procedures

The study protocol dictated that patients from different surgical departments should be enrolled in the study and that these patients should be screened for *S. aureus* colonization prior to enrollment in the study cohort. To achieve this, the local study teams were required to have excellent communication with the participating surgical departments and laboratories. In addition, they had to have the resources to allow for the timely screening of patients. When setting-up this study in different sites across Europe, we encountered that most sites did not have a preoperative *S. aureus* screening policy for surgical patients already in place and that they struggled with implementing a preoperative screening policy. Also, the place and time when patients were seen prior to surgery by a healthcare professional differed quite significantly not only between sites, but also between surgical departments within one site. Because of this, local study teams had a difficult time approaching all eligible patients and arranging that samples were taken in a timely manner, which negatively impacted the full recruitment potential of the hospitals. We overcame these challenges by developing recruitment tools and by applying tailored solutions according to the individual needs of the participating sites.

### Selection of sites

To achieve the proposed sample size of the study cohort, we had to screen a great number of patients for *S. aureus* colonization. Because of budgetary and feasibility reasons, however, we could only select a limited number of sites for participation in the study. This implied that these sites should provide the necessary number of patients for the different surgical procedures for the study. We experienced that several countries did not have many hospitals with a large enough volume of patients to be able to recruit a certain number of subjects for the study. Because of this, the selection of suitable sites for the study took more time than expected. However, by taking the necessary time to select the best sites for the study, we ensured that the study could be executed within appropriate timelines and budget.

In conclusion, despite the challenges, this study will provide important information on the epidemiology of SSI and other infections caused by *S. aureus* in the current surgical population in Europe, thereby supporting the development of future clinical trials of effective interventions and public health interventions aimed at *S. aureus* SSI prevention.

### Future perspectives

There are current efforts to develop novel therapies to combat the increasing problem of antimicrobial resistance. A promising new intervention is antibody-based pre-emptive therapy. A potential advantage of this novel intervention is that it may increase antibiotic effectiveness and may not lead to similar bacterial resistance as antibiotics (14). The current study aims not only to provide contemporary and important information on the incidence, patient-related and contextual factors of SSI and other infections caused by *S. aureus* in the surgical population in Europe, but also to provide data (i.e. sample size and target population) that will directly inform the design of future clinical trials.

## CONCLUSIONS

This prospective observational cohort study on *S. aureus* SSI and postoperative BSI in surgical patients will provide crucial information on the incidence and predictors for this event. Additionally, it will directly impact the design of future clinical trials aimed at *S. aureus* prevention.

## TRIAL STATUS

This manuscript is based on the third version of the ASPIRE-SSI protocol, dated 30 January 2017. The first study cohort subject was enrolled in December 2016, and subject enrollment ended in January 2020.

## Data Availability

The datasets generated and analyzed during the study are not publicly available due to confidentiality reasons but are available from the corresponding author upon scientific review and approval of the request by the Scientific Committee of the study. A summary of the results will be published on clinicaltrials.gov and in peer-reviewed journals.

## LIST OF ABBREVIATIONS

ASA: American Society of Anesthesiologists
ASPIRE-SSI: Advanced Understanding of *Staphylococcus aureus* Infections in Europe - Surgical Site Infections
BSI: bloodstream infection
CDC: Centers for Disease Control and Prevention
COMBACTE: Combatting bacterial resistance in Europe
HAI: healthcare-associated infection
*S. aureus*: *Staphylococcus aureus*
SSI: surgical site infection
SDV: Source data verification.

## DECLARATIONS

### Ethics approval and consent to participate

This study was conducted according to the principles of the Declaration of Helsinki (29), in accordance with the Medical Research Involving Human Subjects Act (WMO) and local guidelines in the participating countries. We received ethics approval of the study protocol, including informed consent procedures, from the Research Ethics Committee or Institutional Review Board of each participating site.

### Consent for publication

Not applicable; this manuscript does not contain any individual person’s data.

### Availability of data and material

The datasets generated and analyzed during the study are not publicly available due to confidentiality reasons but are available from the corresponding author upon scientific review and approval of the request by the Study’s Scientific Committee. A summary of the results will be published on clinicaltrials.gov and in peer-reviewed journals.

### Authors’ contributions

DPRT authored the original study protocol and this manuscript. DPRT, SW, LT, VT, SH, FS, OA and JAJW together created the study design. MW, DH and SW, with help of all other authors designed the Statistical Analysis Plan. All authors read and approved the study protocol and the final manuscript.

## Acknowledgements

We would like to thank the following people for their contributions while writing this study protocol: Mohammed Abbas, Rubana Kalyani, Frank Coenjaerts, Christine Lammens, Edith Schasfoort, and Prof. Surbhi Malhotra-Kumar.

